# Acquired peripheral nerve injuries associated with severe COVID-19

**DOI:** 10.1101/2021.09.24.21263996

**Authors:** Colin K. Franz, Nikhil K. Murthy, George R. Malik, Jean W. Kwak, Dom D’Andrea, Alexis R. Wolfe, Ellen Farr, Melanie A. Stearns, Swati Deshmukh, Jinny O. Tavee, Fang Sun, Kevin N. Swong, Leslie Rydberg, R. James Cotton, Lisa F. Wolfe, James M. Walter, John M. Coleman, John A. Rogers

## Abstract

We diagnosed 66 peripheral nerve injuries in 34 patients who survived severe COVID-19. We combine our latest data with published case series re-analyzed here (117 nerve injuries; 58 patients) to provide a comprehensive accounting of lesion sites. The most common are ulnar (25.1%), common fibular (15.8%), sciatic (13.1%), median (9.8%), brachial plexus (8.7%) and radial (8.2%) nerves at sites known to be vulnerable to mechanical loading. Protection of peripheral nerves should be prioritized in the care of COVID-19 patients. To this end, we report proof of concept data of a wearable, wireless pressure sensor to provide real time monitoring in the intensive care unit setting.

## Introduction

Severe coronavirus disease 2019 (COVID-19) frequently requires intensive care unit (ICU) admission and prolonged periods of mechanical ventilation. ICU acquired weakness (ICU-AW) is an established neuromuscular complication of severe COVID-19^1^ since between 5-17% of patients develop a critical illness from severe acute respiratory syndrome coronavirus 2 infection.^2-4^ ICU-AW clinically manifests as a myopathy, polyneuropathy, or a combination of both.^5^ Regardless, all ICU-AW phenotypes produce diffuse, symmetric symptoms, so if asymmetric neuromuscular symptoms are noted this should trigger further scrutiny for a superimposed process, such as a focal peripheral nerve injury (PNIs).^6, 7^ PNIs may be superimposed on ICU-AW and easily missed in the acute care setting without a detailed neuromuscular assessment. Malik et al. described a case series of COVID-19 PNIs in 12 patients admitted to an inpatient rehabilitation^8^, which has subsequently been reaffirmed by several other published case series in survivors of severe COVID-19.^9-13^ The anatomical distribution of PNIs implies a role for mechanical forces as these localizations mirror sites known to be vulnerable to compression and/or traction.^6, 7^

Unfortunately, the recovery from PNI is notoriously slow and frequently incomplete in the general population^14^, and the demographics of severe COVID-19 patients include enrichment of known risk factors for worse outcomes after PNI such as advanced age, obesity and diabetes mellitus.^8^ There is little doubt that acquired PNIs contribute to long term disability in survivors of severe COVID-19 with reported incidences between 14.5-16%.^8, 9^ Here we use a retrospective chart review to define the types and distribution of PNIs in survivors of severe COVID-19, which is integrated with several smaller published case series.^8-13^ Additionally, we provide feasibility data on a wearable, wireless pressure sensor to provide real time monitoring at the elbow, which has been the single most common nerve compression site in COVID-19 survivors.

## Methods

Study approval was granted by the Northwestern University Institutional Review Board. Patients were identified in retrospective fashion for admissions between April 1, 2020 and March 31, 2021 to 1 of 3 academic medical centers. This included 2 inpatient rehabilitation hospitals (Shirley Ryan AbilityLab, Chicago, IL, USA; Marianjoy Rehabilitation Hospital, Wheaton, IL, USA) and 1 tertiary care hospital (Northwestern Memorial Hospital, Chicago, IL, USA). For the vast majority of PNIs, the diagnosis and localization were supported by electrodiagnostic testing and/or neuromuscular ultrasound (Table 1). We also performed a literature review and included other case series we found (minimum ≥5 patients; Table 2). Due to the retrospective nature of this study, public and patient involvement in this research study design was not obtained.

**Table 1.** Summary of 34 new cases of peripheral nerve injury after severe COVID-19.

| # | Age/Sex | Site | Risk Factors | PNI sites (laterality) | Diagnostics |
| --- | --- | --- | --- | --- | --- |
| 1 | 60s/M | SRAlab | DM, Obese | Sciatic (left), Fibular (right) | EDX (axonal) |
| 2 | 50s/M | SRAlab | DM | Ulnar (left) | EDX (demyelin.) |
| 3 | 40s/F | SRAlab | Obese | Cervical radiculopathy (left) | EDX (axonal) |
| 4 | 60s/M | SRAlab | DM | Femoral (left), Obturator (left), Spinal accessory (right) | EDX (axonal), MRI (pelvis) |
| 5 | 70s/M | SRAlab | None | Sciatic (right), Fibular (left) | EDX (axonal) |
| 6 | 30s/M | SRAlab | Obese | Sciatic (right), Fibular (left), Phrenic (right) | EDX (axonal), US (phrenic) |
| 7 | 40s/M | SRAlab | None | Phrenic (right) | US (phrenic) |
| 8 | 60s/M | SRAlab | HIV | Radial (right), Ulnar (right), Median (right), Sciatic (right), Ulnar (left), Median (left) | EDX (axonal; but demyelin. median) |
| 9 | 60s/M | SRAlab | Obese | Brachial plexopathy (right), Ulnar (right), Median (right), Phrenic (right) | EDX (axonal; but demyelin. ulnar), US (phrenic) |
| 10 | 60s/M | SRAlab | None | Fibular (right) | EDX (axonal) |
| 11 | 70s/M | SRAlab | CKD | Femoral (right), Obturator (right), Fibular (right) | EDX (axonal), MRI (pelvis) |
| 12 | 50s/M | SRAlab | DM | Cervical radiculopathy (left), Ulnar (left), Phrenic (right) | EDX (axonal), MRI (neck/shoulder), US (phrenic) |
| 13 | 60s/M | SRAlab | DM | Sciatic (right), Phrenic (right) | EDX (axonal), US (phrenic) |
| 14 | 30s/M | SRAlab | Obesity | Sciatic (left) | EDX (axonal) |
| 15 | 40s/M | SRAlab | None | Fibular (left), Fibular (right) | EDX (axonal) |
| 16 | 60s/M | SRAlab | None | Ulnar (right), Radial (right), Median (right), Ulnar (left), Radial (left) | EDX (axonal) |
| 17 | 70s/F | SRAlab | DM | Sciatic (left) | EDX (axonal) |
| 18 | 70s/M | SRAlab | DM, CKD | Fibular (left) | EDX (axonal) |
| 19 | 50s/M | SRAlab | Obese | Sciatic (right), Fibular (left) | EDX (axonal) |
| 20 | 40s/M | SRAlab | DM, Obese | Fibular (right), Fibular (left) | EDX (axonal) |
| 21 | 70s/M | SRAlab | None | Ulnar (left), Lateral femoral cutaneous (left) | EDX (axonal, but clinical LFCN) |
| 22 | 50s/M | SRAlab | None | Ulnar (left) | EDX (axonal) |
| 23 | 50s/M | NMH | DM, Obese | Radial (right) | None (clinical) |
| 24 | 30s/F | NMH | DM, Obese | Sciatic (left) | EDX (axonal) |
| 25 | 30s/M | NMH | Obese | Ulnar (left) | EDX (axonal) |
| 26 | 40s/M | NMH | Obese | Radial (left) | EDX (axonal) |
| 27 | 20s/M | NMH | DM, Obese | Ulnar (left) | EDX (axonal) |
| 28 | 20s/M | NMH | Obese | Brachial plexopathy (left), Radial (right) | EDX (axonal) |
| 29 | 60s/M | NMH | Obese | Brachial plexopathy (left), Ulnar (left), Sciatic (left), Fibular (right) | EDX (axonal; but demyelin. ulnar) |
| 30 | 30s/F | MRH | DM, Obese | Fibular (right), Fibular (left) | EDX (axonal) |
| 31 | 60s/M | MRH | None | Fibular (right) | EDX (axonal) |
| 32 | 60s/M | MRH | DM, Obese | Brachial plexopathy (right) | EDX (axonal) |
| 33 | 70s/M | MRH | DM, Obese | Fibular (right), Fibular (left) | None (clinical) |
| 34 | 30s/M | MRH | DM, Obese | Median (right), Median (left) | EDX (axonal) |

PNI, peripheral nerve injury; SRAlab, Shirley Ryan AbilityLab (rehabilitation hospital); NMH, Northwestern Memorial Hospital; MJH, Marianjoy Rehabilitation Hospital; DM, diabetes mellitus; HIV, human immunodeficiency
virus; CKD, chronic kidney disease; EDX, electrodiagnostics; MRI, magnetic resonance imaging; US, ultrasound; demyelin., demyelinating; LFCN, lateral femoral cutaneous nerve. In accordance with journal policy, patient age is expressed according to their decade of life.

In addition, proof of concept study of wireless, soft, skin-interfaced pressure sensor^15, 16^ was performed on 2 patients admitted to ICU for severe COVID-19. This technology, comprised of a Bluetooth communication system, enables real-time, continuous, and wireless monitoring of pressure, through a smartphone (weight: ∼4 g; dimensions: ∼6.5 cm x 2 cm x 0.4 cm (length x width x height); Figure 2A-B). After following a sterilization process, deployment of the sensor with a thin adhesive dressing (Tegaderm, 3M) secured the intimate contact with the skin on the elbow (Figure 2C). The sensor was applied adjacent to the ulnar nerve onto the medial epicondyle, without interfering with clinical standard-of-care equipment for the COVID-19 treatment. The wireless connection to a smartphone allowed for real-time visual display through a graphical user interface, data transmission, collection, and storage.

## Results

The baseline characteristics of 34 patients that acquired nerve injury after surviving severe COVID-19 are reported in Table 1. Of those patients, 4 are female and 30 are male, with an average age of 53 ± 15 years (range 21-77) and average body mass index (BMI) of 33 ± 10 (range 17-59). Sixteen patients carry a diagnosis of diabetes mellitus (47%), and a total of 19 patients are classified as obese (BMI>30; 56%). There are a total of 66 PNIs described here, which makes for an average of 1.9 ± 1.2 PNIs (range 1-6) per patient in our population. There are 12 different nerve injury sites in this case series, with the most common localizations being the common fibular (n=16), ulnar (n=12), sciatic (n=9), radial (n=6), and median (n=6). Multifocal PNIs (≥2 sites) were noted in 53% (n=18) of the cases. Electrodiagnostic testing was available for 57 of these PNIs, and 93% (n=53) show electrophysiological evidence of more severe axonotmesis grade injury as compared to 7% (n=4) with lesser neuropraxia (demyelinating) grade^17^.

For the subset of these cases from the Shirley Ryan AbilityLab rehabilitation hospital (n=22), as well as additional published cases from the same site (n=12)^8^, we calculated that these represented 10.7% (34 of 319) of the post-COVID-19 admissions over a 12 month period. All of the new cases except for one (case #7; Table 1) had PNI diagnosed in the electrodiagnostic testing laboratory. In contrast, out of the non-COVID-19 related general medical rehabilitation admissions over the same period only 0.5% (3 of 572) had a PNI diagnosed in the electrodiagnostic testing laboratory. For the remaining admissions to this site, the majority of which included traumatic and non-traumatic disorders of the central nervous system, only 0.6% (13 of 2122) had a PNI diagnosed in the electrodiagnostic testing laboratory.

When our current data is combined with prior case series of PNIs in survivors of COVID-19 (Table 2), there are 183 PNIs, at 16 different sites in 92 patients. The most common localizations overall are ulnar (25.1%), common fibular (15.8%), sciatic (13.1%), median (9.8%), brachial plexus (8.7%) and radial nerve (8.2%), which is graphically summarized in Figure 1. In several of the cases we report here we noted skin pressure sores overlaying the nerve compression sites including the fibular head and elbow. This was the inspiration for us to test the feasibility of wireless pressure sensor on COVID patients admitted to the ICU. Figures 1D and E illustrate the case of a patient being treated with high flow oxygen via nasal cannula and self proning for COVID-19 pneumonia. The subject was conscious during the protocol, which allowed for self-adjustments of the arms. The pressures from both left and right elbows at the medial epicondyle (Figures 1F-G) show similar values, the averages of which are 44.9 mmHg and 41.7 mmHg, respectively. The inconsistency over time suggests that the subject repositioned the arms, which is also shown in the varying standard deviations, 11.0 mmHg and 3.2 mmHg, respectively. Figure 2H shows the photograph of an intubated patient with respiratory failure from COVID-19 pneumonia, who was in a swimmer’s position and in a reverse Trendelenburg position with chest paddings (Figure 2G). This variation of the prone position can be used to protect the ulnar nerve at the medial elbow^18^ and reduces the pressure reading to 4.5 ± 0.6 mmHg (Figure 2I). Gentle reloading of the medial elbow by the placement of a cushion underneath it increases the pressure to 17.2 ± 1.0 mmHg as detected by the wireless pressure sensor (Figure 2I).

**Figure 1.**
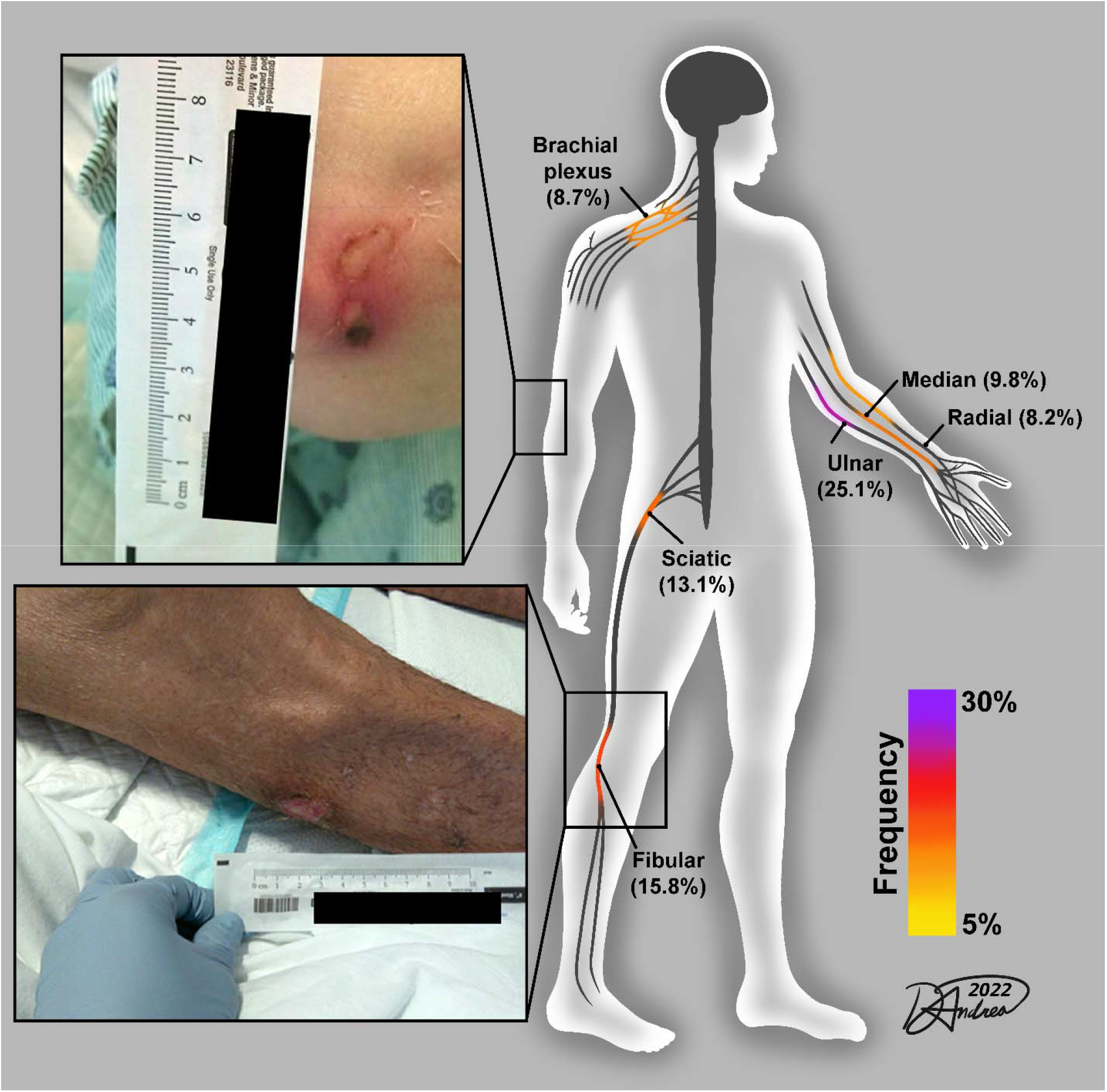
Graphical summary of common nerve injury sites in survivors of COVID-19. This summary includes current data and several recent case series of peripheral nerve injuries associated with COVID-19. Inset photographs show examples skin pressure sores overlaying the nerve compression sites – elbow (top) and fibular head (bottom).

**Figure 2.**
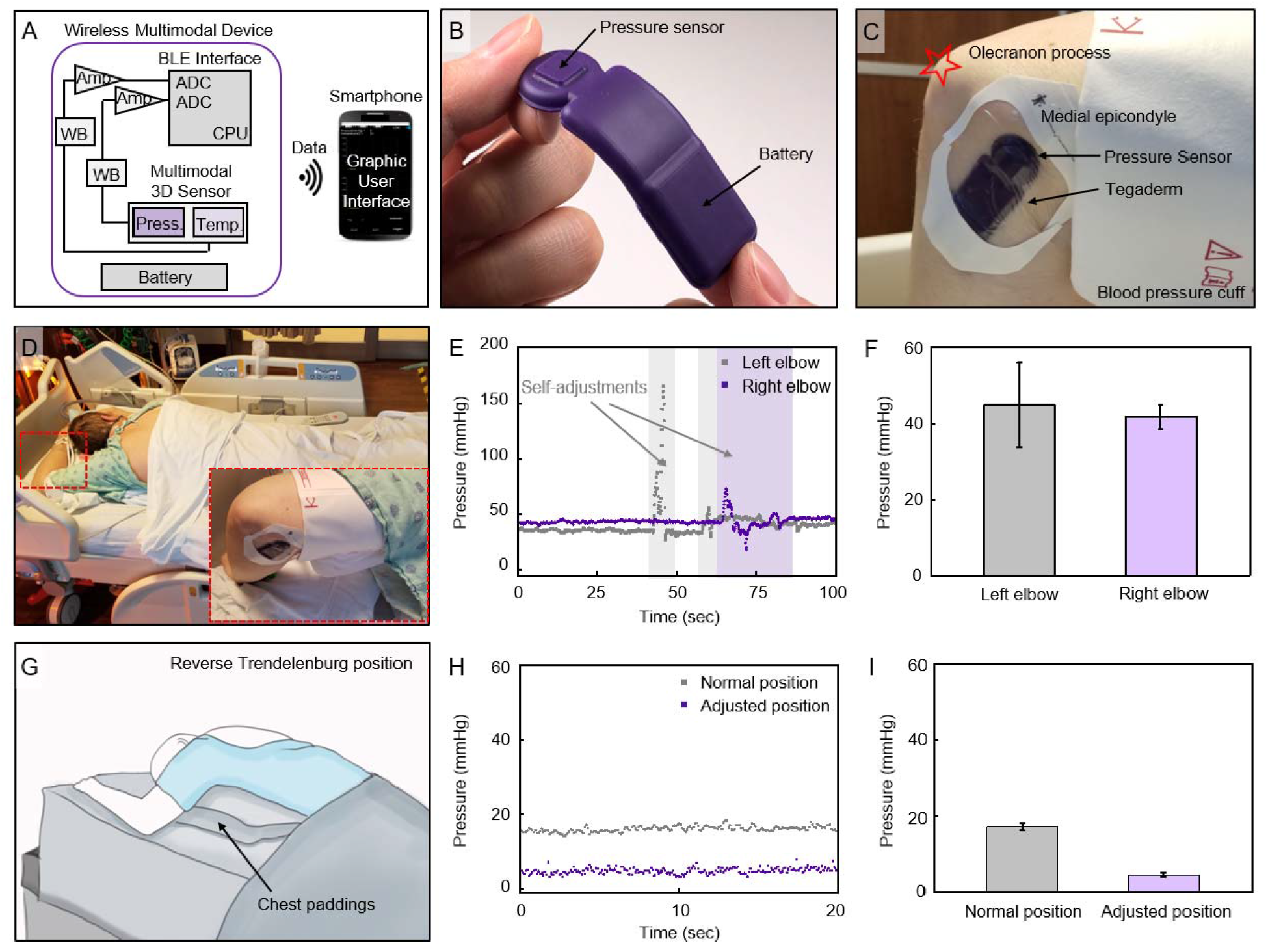
Soft, skin-interfaced sensor for wireless measurements of pressure for COVID19 patients in ICU. (A) Functional block diagram of the system that is powered through a battery, illustrating a Bluetooth Low Energy (BLE) system-on-a-chip (SoC), which connects to a Wheatstone bridge (WB) and an instrumentation amplifier (Amp) that convert and amplify the signal of the pressure sensor. The analog-to-digital converter (ADC)-sampled data passes through the central processing unit (CPU), which then transmits to BLE radio, displaying real-time data on the graphical user interface (smartphone). (B) Photograph of the sensor, depicting its thin and flexible form factor. (C) Photograph of a subject’s elbow, wearing the Tegaderm-secured sensor at the medial epicondyle. (G) Illustration of an intubated subject with the left arm up, in a reverse Trendelenburg position with chest paddings. (E) Representative pressure data of the self-proning subject at both left and right arms. Colored regions indicate self-adjustments. (F) Pressure measured in demonstrated normal prone position on both arms (measured at 10 Hz for 200, 300 seconds, respectively, data with individual sensor; error bar: SD). (D) Photograph of a self-proning subject (i.e. patient rolls themselves over on belly), placing his arms with the sensors upward. Red-dashed rectangle shows the unloading of the arm. (H) Representative pressure data of an intubated subject in the demonstrated normal prone position and the adjusted/optimized position. (I) Pressure measured in demonstrated normal prone position and adjusted, optimized prone position (measured at 10 Hz for 60, 900 seconds, respectively, data with the same sensor; error bar: SD).

## Discussion

We report on a surprisingly large case series of 34 patients with 66 PNIs associated with survival from severe COVID-19. In majority of these cases there are ≥2 PNIs affecting the same person (Table 1). When taken in the context of our literature review of nerve injury case series associated with COVID-19 (n=5-15 patients/study), these data show acquired PNIs are an important contributor to prolonged neurological impairments in survivors of COVID-19. The mechanisms underlying the propensity for PNI in COVID-19 critical illness is difficult to establish, but the anatomical distribution (Figure 1) of these injuries implicate mechanical forces such as pressure against bony prominence.^19^ In a proof of concept study we demonstrate the feasibility of a wearable, wireless pressure sensor system to provide real time monitoring at the medial elbow (Figure 2), which is most common site of compressive neuropathy in severe COVID-19 (Figure 1).

The risk for focal PNIs in critically ill patients is well known^20^, but the incidence has not been defined. In part this may be because these injuries may overlap with ICU-AW and not get the dedicated attention needed to diagnose with imaging^7^ or electrodiagnostics^5^. In certain cases, like phrenic nerve injuries, the best diagnostic option may be a neuromuscular ultrasound study^21^ but not all hospital systems have access to such expertise. Occasionally nerve compression in severe COVID-19 survivors may be accounted for by a hematoma^7^ or an iatrogenic cause such as focal neuritis adjacent to central line site^21^ that can be diagnosed by advanced imaging modalities.

Early results in severe COVID-19 survivors from single center case series put the incidence reported incidences between 14.5-16%.^8, 9^ In the present report we report an incidence for a subset of the patients admitted to a single rehabilitation center of 10.7%, which is just slightly lower than prior reports. There were high rates of diabetes mellitus, obesity, male sex, and older age seen in our cohort which are characteristics of severe COVID-19-related ARDS patients^2^, and risk factors for PNI in these patients.^19, 22^ On a cellular level, a combination of inflammatory and immune-mediated injury caused by COVID-19 may increase susceptibility to nerve injury when patients have severe disease.^23^ There is a paucity of evidence for direct SARS-CoV-2 of peripheral nerves, but this can’t be dismissed as a factor in a small subset of cases.^24^

Our use of wireless pressure sensors to monitor areas of ulnar nerve compression demonstrates a future approach to preventing prone positioning-related nerve injury. The sensors can provide real-time pressure information and can be used to adjust positioning at known compression sites before a compressive neuropathy occurs, while being used for extended periods of time (Figure 2). Patients with severe COVID-19 appear particularly susceptible to positioning related PNIs.^6, 7^ For example, the prone positioning intervention has been recommended for 12 to 16 hours per day in mechanically ventilated adults with COVID-19 and refractory hypoxemia^25^, but has associated with increase rates of acquired peripheral PNIs.^6-8, 10-12^

Limitations of this study include lack of a control group, and the retrospective design, which precludes establishment of a causal relationship between patient positioning and peripheral nerve injury. Additionally, some patients with less severe PNIs may not have had advanced imaging or electrodiagnostic studies ordered, which may have led to a bias towards more severe PNIs and underestimation of PNI incidence.

Given the ongoing COVID-19 pandemic, and risks of new variants causing a resurgence of hospital admissions, further attention should be paid to the long-term sequela including PNIs. These injuries require long-term follow up and care with therapy and rehabilitation. Prevention and early identification of these injuries could help decrease additional morbidity of the disease.

## Data Availability

Data will be made available upon request.

## Acknowledgements

We would like to acknowledge Dr. Richard G Wunderink, MD (Professor, Pulmonary and Critical Care, Northwestern Feinberg School of Medicine) and Helen Donnelly, RN (Clinical Research Nurse, Pulmonary and Critical Care, Northwestern Feinberg School of Medicine) for assisting us with patient recruitment for our pressure sensor studies. We would like to acknowledge Peggy Kirk, RN (President and Chief Executive Officer, Shirley Ryan AbilityLab) for supporting this project as a part of a continuous quality improvement to enhance patient safety and outcomes.

---

**Supplemental Table 2.**
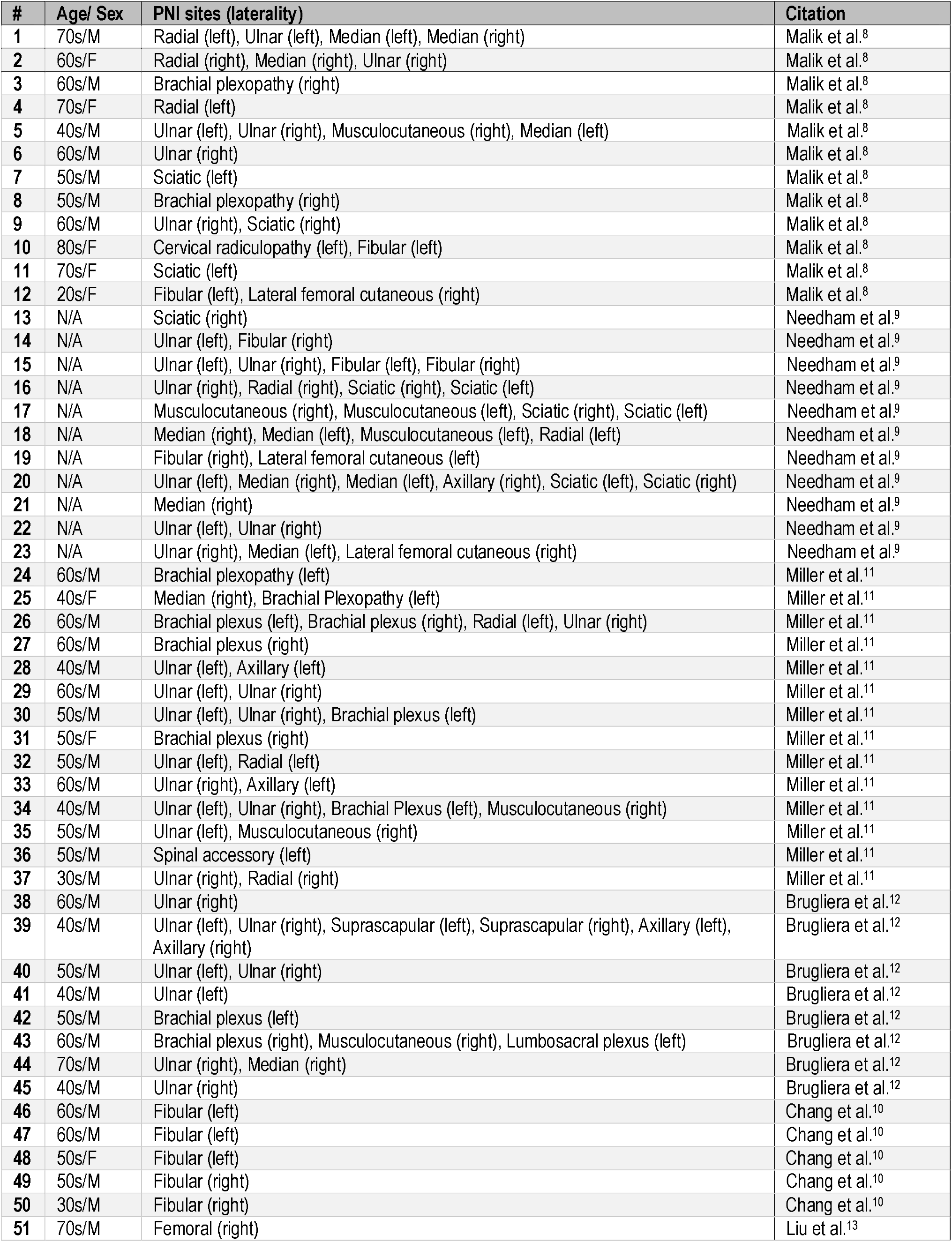
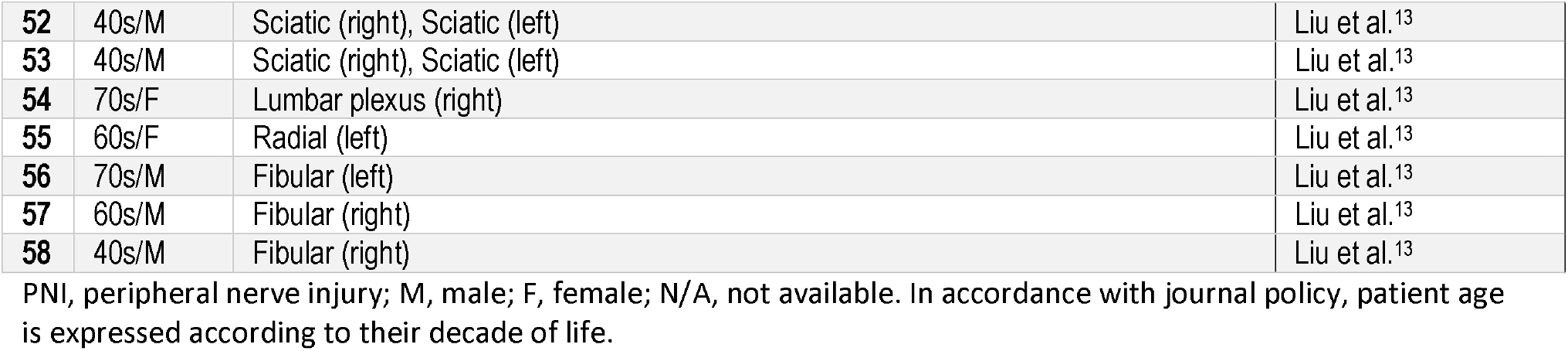
Summary of patient characteristics from previously published case series.

| # | Age/ Sex | PNI sites (laterality) | Citation |
| --- | --- | --- | --- |
| 1 | 70s/M | Radial (left), Ulnar (left), Median (left), Median (right) | Malik et al.<sup>8</sup> |
| 2 | 60s/F | Radial (right), Median (right), Ulnar (right) | Malik et al.<sup>8</sup> |
| 3 | 60s/M | Brachial plexopathy (right) | Malik et al.<sup>8</sup> |
| 4 | 70s/F | Radial (left) | Malik et al.<sup>8</sup> |
| 5 | 40s/M | Ulnar (left), Ulnar (right), Musculocutaneous (right), Median (left) | Malik et al.<sup>8</sup> |
| 6 | 60s/M | Ulnar (right) | Malik et al.<sup>8</sup> |
| 7 | 50s/M | Sciatic (left) | Malik et al.<sup>8</sup> |
| 8 | 50s/M | Brachial plexopathy (right) | Malik et al.<sup>8</sup> |
| 9 | 60s/M | Ulnar (right), Sciatic (right) | Malik et al.<sup>8</sup> |
| 10 | 80s/F | Cervical radiculopathy (left), Fibular (left) | Malik et al.<sup>8</sup> |
| 11 | 70s/F | Sciatic (left) | Malik et al.<sup>8</sup> |
| 12 | 20s/F | Fibular (left), Lateral femoral cutaneous (right) | Malik et al.<sup>8</sup> |
| 13 | N/A | Sciatic (right) | Needham et al.<sup>9</sup> |
| 14 | N/A | Ulnar (left), Fibular (right) | Needham et al.<sup>9</sup> |
| 15 | N/A | Ulnar (left), Ulnar (right), Fibular (left), Fibular (right) | Needham et al.<sup>9</sup> |
| 16 | N/A | Ulnar (right), Radial (right), Sciatic (right), Sciatic (left) | Needham et al.<sup>9</sup> |
| 17 | N/A | Musculocutaneous (right), Musculocutaneous (left), Sciatic (right), Sciatic (left) | Needham et al.<sup>9</sup> |
| 18 | N/A | Median (right), Median (left), Musculocutaneous (left), Radial (left) | Needham et al.<sup>9</sup> |
| 19 | N/A | Fibular (right), Lateral femoral cutaneous (left) | Needham et al.<sup>9</sup> |
| 20 | N/A | Ulnar (left), Median (right), Median (left), Axillary (right), Sciatic (left), Sciatic (right) | Needham et al.<sup>9</sup> |
| 21 | N/A | Median (right) | Needham et al.<sup>9</sup> |
| 22 | N/A | Ulnar (left), Ulnar (right) | Needham et al.<sup>9</sup> |
| 23 | N/A | Ulnar (right), Median (left), Lateral femoral cutaneous (right) | Needham et al.<sup>9</sup> |
| 24 | 60s/M | Brachial plexopathy (left) | Miller et al.<sup>11</sup> |
| 25 | 40s/F | Median (right), Brachial Plexopathy (left) | Miller et al.<sup>11</sup> |
| 26 | 60s/M | Brachial plexus (left), Brachial plexus (right), Radial (left), Ulnar (right) | Miller et al.<sup>11</sup> |
| 27 | 60s/M | Brachial plexus (right) | Miller et al.<sup>11</sup> |
| 28 | 40s/M | Ulnar (left), Axillary (left) | Miller et al.<sup>11</sup> |
| 29 | 60s/M | Ulnar (left), Ulnar (right) | Miller et al.<sup>11</sup> |
| 30 | 50s/M | Ulnar (left), Ulnar (right), Brachial plexus (left) | Miller et al.<sup>11</sup> |
| 31 | 50s/F | Brachial plexus (right) | Miller et al.<sup>11</sup> |
| 32 | 50s/M | Ulnar (left), Radial (left) | Miller et al.<sup>11</sup> |
| 33 | 60s/M | Ulnar (right), Axillary (left) | Miller et al.<sup>11</sup> |
| 34 | 40s/M | Ulnar (left), Ulnar (right), Brachial Plexus (left), Musculocutaneous (right) | Miller et al.<sup>11</sup> |
| 35 | 50s/M | Ulnar (left), Musculocutaneous (right) | Miller et al.<sup>11</sup> |
| 36 | 50s/M | Spinal accessory (left) | Miller et al.<sup>11</sup> |
| 37 | 30s/M | Ulnar (right), Radial (right) | Miller et al.<sup>11</sup> |
| 38 | 60s/M | Ulnar (right) | Brugliera et al.<sup>12</sup> |
| 39 | 40s/M | Ulnar (left), Ulnar (right), Suprascapular (left), Suprascapular (right), Axillary (left), Axillary (right) | Brugliera et al.<sup>12</sup> |
| 40 | 50s/M | Ulnar (left), Ulnar (right) | Brugliera et al.<sup>12</sup> |
| 41 | 40s/M | Ulnar (left) | Brugliera et al.<sup>12</sup> |
| 42 | 50s/M | Brachial plexus (left) | Brugliera et al.<sup>12</sup> |
| 43 | 60s/M | Brachial plexus (right), Musculocutaneous (right), Lumbosacral plexus (left) | Brugliera et al.<sup>12</sup> |
| 44 | 70s/M | Ulnar (right), Median (right) | Brugliera et al.<sup>12</sup> |
| 45 | 40s/M | Ulnar (right) | Brugliera et al.<sup>12</sup> |
| 46 | 60s/M | Fibular (left) | Chang et al.<sup>10</sup> |
| 47 | 60s/M | Fibular (left) | Chang et al.<sup>10</sup> |
| 48 | 50s/F | Fibular (left) | Chang et al.<sup>10</sup> |
| 49 | 50s/M | Fibular (right) | Chang et al.<sup>10</sup> |
| 50 | 30s/M | Fibular (right) | Chang et al.<sup>10</sup> |
| 51 | 70s/M | Femoral (right) | Liu et al.<sup>13</sup> |
| 52 | 40s/M | Sciatic (right), Sciatic (left) | Liu et al.<sup>13</sup> |
| 53 | 40s/M | Sciatic (right), Sciatic (left) | Liu et al.<sup>13</sup> |
| 54 | 70s/F | Lumbar plexus (right) | Liu et al.<sup>13</sup> |
| 55 | 60s/F | Radial (left) | Liu et al.<sup>13</sup> |
| 56 | 70s/M | Fibular (left) | Liu et al.<sup>13</sup> |
| 57 | 60s/M | Fibular (right) | Liu et al.<sup>13</sup> |
| 58 | 40s/M | Fibular (right) | Liu et al.<sup>13</sup> |

PNI, peripheral nerve injury; M, male; F, female; N/A, not available. In accordance with journal policy, patient age is expressed according to their decade of life.

## References

1. Suh J, Amato AA. Neuromuscular complications of coronavirus disease-19. Curr Opin Neurol. 2021 Jun 21.

2. Fried MW, Crawford JM, Mospan AR, et al. Patient Characteristics and Outcomes of 11 721 Patients With Coronavirus Disease 2019 (COVID-19) Hospitalized Across the United States. Clin Infect Dis. 2021 May 18;72(10):e558–e65.

3. Grasselli G, Zangrillo A, Zanella A, et al. Baseline Characteristics and Outcomes of 1591 Patients Infected With SARS-CoV-2 Admitted to ICUs of the Lombardy Region, Italy. JAMA. 2020 Apr 28;323(16):1574–81.

4. Wu Z, McGoogan JM. Characteristics of and Important Lessons From the Coronavirus Disease 2019 (COVID-19) Outbreak in China: Summary of a Report of 72314 Cases From the Chinese Center for Disease Control and Prevention. JAMA. 2020 Apr 7;323(13):1239–42.

5. Koshy K, Zochodne DW. Neuromuscular complications of critical illness. Handb Clin Neurol. 2013;115:759–80.

6. Simpson AI, Vaghela KR, Brown H, et al. Reducing the Risk and Impact of Brachial Plexus Injury Sustained From Prone Positioning-A Clinical Commentary. J Intensive Care Med. 2020 Dec;35(12):1576–82.

7. Fernandez CE, Franz CK, Ko JH, et al. Imaging Review of Peripheral Nerve Injuries in Patients with COVID-19. Radiology. 2021 Mar;298(3):E117–E30.

8. Malik GR, Wolfe AR, Soriano R, et al. Injury-prone: peripheral nerve injuries associated with prone positioning for COVID-19-related acute respiratory distress syndrome. Br J Anaesth. 2020 Dec;125(6):e478–e80.

9. Needham E, Newcombe V, Michell A, et al. Mononeuritis multiplex: an unexpectedly frequent feature of severe COVID-19. J Neurol. 2021 Aug;268(8):2685–9.

10. Chang LG, Zar S, Seidel B, Kurra A, Gitkind A. COVID-19 Proned Ventilation and Its Possible Association With Foot Drop: A Case Series. Cureus. 2021 Apr 8;13(4):e14374.

11. Miller C, O’Sullivan J, Jeffrey J, Power D. Brachial Plexus Neuropathies During the COVID-19 Pandemic: A Retrospective Case Series of 15 Patients in Critical Care. Phys Ther. 2021 Jan 4;101(1).

12. Brugliera L, Filippi M, Del Carro U, et al. Nerve Compression Injuries After Prolonged Prone Position Ventilation in Patients With SARS-CoV-2: A Case Series. Arch Phys Med Rehabil. 2021 Mar;102(3):359–62.

13. Liu EA, Salazar T, Chiu E, et al. Focal Peripheral Neuropathies Observed in Patients Diagnosed With COVID-19: A Case Series. Am J Phys Med Rehabil. 2022 Feb 1;101(2):164–9.

14. Lundborg G. A 25-year perspective of peripheral nerve surgery: evolving neuroscientific concepts and clinical significance. J Hand Surg Am. 2000 May;25(3):391–414.

15. Kwak JW, Han M, Xie Z, et al. Wireless sensors for continuous, multimodal measurements at the skin interface with lower limb prostheses. Sci Transl Med. 2020 Dec 16;12(574).

16. Park Y, Kwon K, Kwak SS, et al. Wireless, skin-interfaced sensors for compression therapy. Sci Adv. 2020 Dec;6(49).

17. Seddon HJ. A Classification of Nerve Injuries. Br Med J. 1942 Aug 29;2(4260):237–9.

18. Lee CT, Espley AJ. Perioperative ulnar neuropathy in orthopaedics: association with tilting the patient. Clin Orthop Relat Res. 2002 Mar(396):106–11.

19. Practice Advisory for the Prevention of Perioperative Peripheral Neuropathies 2018: An Updated Report by the American Society of Anesthesiologists Task Force on Prevention of Perioperative Peripheral Neuropathies. Anesthesiology. 2018 Jan;128(1):11–26.

20. Angel MJ, Bril V, Shannon P, Herridge MS. Neuromuscular function in survivors of the acute respiratory distress syndrome. Can J Neurol Sci. 2007 Nov;34(4):427–32.

21. Patel Z, Franz CK, Bharat A, et al. Diaphragm and Phrenic Nerve Ultrasound in COVID-19 Patients and Beyond: Imaging Technique, Findings, and Clinical Applications. J Ultrasound Med. 2021 Mar 27.

22. Warner MA, Warner ME, Martin JT. Ulnar neuropathy. Incidence, outcome, and risk factors in sedated or anesthetized patients. Anesthesiology. 1994 Dec;81(6):1332–40.

23. Suh J, Mukerji SS, Collens SI, et al. Skeletal Muscle and Peripheral Nerve Histopathology in COVID-19. Neurology. 2021 Aug 24;97(8):e849–e58.

24. Matschke J, Lutgehetmann M, Hagel C, et al. Neuropathology of patients with COVID-19 in Germany: a post-mortem case series. Lancet Neurol. 2020 Nov;19(11):919–29.

25. Care of Critically Ill Adult Patients With COVID-19: National Institutes of Health2021 9/13/2021.

